# Gut microbiota composition of stunted children in Banyumas Regency, Indonesia: A 16S rRNA metagenomic analysis

**DOI:** 10.64898/2025.12.07.25341778

**Authors:** Rizqi Yanuar Pauzi, Suci Ihtiaringtyas, Novia Yunika, Glory Aprilia Kusumawardani

**Affiliations:** Department of Microbiology, Faculty of Medicine, Universitas Jenderal Soedirman, Purwokerto 53147, Indonesia; Department of Parasitology, Faculty of Medicine, Universitas Jenderal Soedirman, Purwokerto 53147, Indonesia; Faculty of Medicine, Universitas Jenderal Soedirman, Purwokerto 53147, Indonesia

**Keywords:** dysbiosis, gut microbiota, nutritional intake, stunting, 16S rRNA sequencing

## Abstract

Stunting remains a major public health challenge in Indonesia and is increasingly associated with gut microbiota dysbiosis. This study examined 36 children aged 2–5 years through anthropometry, dietary assessment, and pooled fecal sampling. Full-length 16S rRNA sequencing using Oxford Nanopore Technologies revealed notable microbial alterations in the stunted group. Stunted children exhibited reduced alpha diversity and lower microbial richness, indicating a simplified gut ecosystem. Although both groups were dominated by Bacillota (>96%), stunted children showed higher proportions of Clostridia affiliated orders, including Eubacteriales, Peptostreptococcales, and Erysipelotrichaceae, along with enrichment of fermentative and dysbiosis associated genera such as Blautia, Romboutsia, and Terrisporobacter. Beneficial fiber degrading taxa, including Lachnospiraceae, were proportionally higher in normal children. Functional predictions using PICRUSt2 revealed greater microbial metabolic activity in the stunting group, particularly in carbohydrate, amino acid, and nucleotide metabolism, with elevated pathways such as starch and sucrose metabolism, glycolysis, and porphyrin and pyrimidine metabolism. Dietary assessment showed significantly lower intake of energy, protein, fat, and multiple micronutrients among stunted children, consistent with observed microbial and functional alterations. These findings indicate a distinct fermentative dysbiosis in Banyumas stunted children and highlight the need for integrated nutritional and microbiota targeted interventions.

## Introduction

Stunting, a chronic condition resulting from prolonged undernutrition, affects more than 150 million children under the age of five globally and remains a major public health concern, particularly in Southeast Asia (World Health Organization 2023). In Indonesia, the prevalence of stunting was 21.5% in 2023, a figure that remains well above the World Health Organization targets and underscores persistent challenges in early childhood nutrition and health (Kemenkes 2023). Stunting is associated with long-term adverse outcomes, including impaired cognitive development, reduced physical capacity, and an increased risk of metabolic and cardiovascular diseases later in life (Pauzi et al. 2025).

The pathophysiology of stunting is multifactorial, extending beyond inadequate dietary intake to include recurrent infections and environmental enteric dysfunction (EED), a subclinical condition of the small intestine characterized by villous atrophy, increased permeability, and persistent inflammation (Budge et al. 2019). Emerging evidence strongly implicates the gut microbiota, the complex community of microorganisms residing in the gastrointestinal tract, as a central regulator of nutrient metabolism, immune system maturation, and gut barrier integrity (Ronan et al. 2021). An imbalance in this microbial ecosystem, termed dysbiosis, has been linked to the pathogenesis of EED and subsequent growth faltering (Amimo et al. 2025). Experimental studies further demonstrate that dysbiosis, when combined with undernutrition, can induce EED-like phenotypes and significantly impair growth, suggesting a bidirectional interaction between microbial imbalance and intestinal dysfunction (Perruzza et al. 2024).

Studies examining the gut microbiota of stunted children have reported heterogeneous findings across different geographic regions. In Indonesia, investigations conducted in West Sumatra, Banten, and West Java have identified variations in the relative abundances of key bacterial phyla such as Firmicutes, Proteobacteria, and Bacteroidetes, as well as differences in the prevalence of genera such as Prevotella (Masrul et al. 2020; Surono et al. 2021). These inconsistencies likely reflect regional differences in diet, environmental exposures, host genetics, and sociocultural practices. Such variation highlights the importance of conducting localized microbiome studies to capture population-specific microbial signatures (Kandpal et al. 2022).

Banyumas Regency in Central Java represents an area with a persistently high risk of stunting and documented challenges in sanitation and clean water access, factors known to influence gut microbial composition (Dinkes 2023). Despite this, no prior studies have described the gut microbiome of children in this region. To address this gap, the present study provides the first comprehensive characterization of the gut microbiota in stunted and normal-weight children in Banyumas using 16S rRNA gene sequencing. We hypothesized that stunted children would exhibit reduced microbial diversity, enrichment of pro-inflammatory taxa, and altered functional metabolic pathways compared with their normal-weight peers, reflecting a dysbiotic microbiome that may contribute to growth impairment.

## Materials And Methods

### Study Design and Participants

A cross-sectional study was conducted in the Tambak district of Banyumas Regency, Indonesia, from June to September 2025. A total of 36 children aged 2–5 years were recruited using purposive and quota sampling to obtain two groups: 18 children with stunting and 18 with normal nutritional status. Stunting was defined as a height-for-age Z-score (HAZ) < –2, whereas normal nutritional status corresponded to HAZ values between –2 and +3, in accordance with the WHO Child Growth Standards. Children who had received antibiotics within the previous month or who presented with acute gastrointestinal infections were excluded. The study was approved by the Health Research Ethics Committee of the Faculty of Medicine, Jenderal Soedirman University. Written informed consent was obtained from all parents or legal guardians prior to participation.

### Data and Sample Collection

Anthropometric measurements, including height, weight, head circumference, and mid-upper arm circumference, were obtained by trained personnel using standardized procedures. Height was measured using a stadiometer or infantometer to the nearest 0.1 cm, and body weight was recorded with a calibrated digital scale to the nearest 0.1 kg. HAZ, WAZ, and WHZ indices were calculated to assess nutritional status. Sociodemographic information, health history, and dietary intake were collected using structured questionnaires. Dietary intake was assessed with a Semi-Quantitative Food Frequency Questionnaire (SQ-FFQ) to estimate macronutrient and micronutrient consumption.

For microbiome analysis, stool samples were collected in sterile containers provided to guardians along with detailed instructions. Samples were collected in the morning, immediately stored in a cooler box with ice packs (2–8°C), and transported to the field laboratory within 4 hours. Upon arrival, samples were stored at −80°C until DNA extraction. For sequencing purposes, stool samples from stunted children were pooled into a single composite sample, and likewise, stool samples from children with normal nutritional status were pooled, resulting in two final samples representing each group. Pooling was performed to increase sequencing depth per group and because this study was designed as an exploratory pilot analysis.

### DNA Extraction and 16S rRNA Gene Sequencing

Total microbial genomic DNA was extracted from 250 mg of each fecal sample. Prior to extraction, stool samples were homogenized using a sterile swab stick. Approximately 250 mg of homogenized material was transferred into a 1.5 mL microtube containing 150 µL of PBS and vortexed at 1200 rpm for 3 minutes. A 350 µL aliquot of the homogenate was processed using the Quick-DNA™ Fecal/Soil Microbe Kit (D6010, Zymo Research) following the manufacturer’s protocol. DNA concentration was quantified using a Qubit 4 Fluorometer (Invitrogen), and purity was assessed using a Nanophotometer N50 (Implen). Samples with DNA concentrations ≥ 3 ng/µL were used for library preparation.

Full-length 16S rRNA genes were amplified using the primer sets provided in the 16S Barcoding Kit V14 (SQK-16S114.24, Oxford Nanopore Technologies), targeting the entire ∼1500 bp gene and enabling sample differentiation through up to 24 barcodes. Barcoding, amplification, and purification were performed according to the manufacturer’s instructions. Sequencing libraries were prepared using the 16S Barcoding Kit V14 workflow, pooled, and loaded onto a MinION Mk1D device equipped with an R10.4.1 flow cell (Oxford Nanopore Technologies). Sequencing was performed according to the standard MinION protocol.

### Bioinformatic and Statistical Analysis

Raw sequencing data generated by the Oxford Nanopore Technologies (ONT) platform were basecalled using Guppy v6.5.7 in super-accuracy mode. Trimmed and demultiplexed FASTQ reads were analyzed using the wf-metagenomics workflow with a minimum read length threshold of 1000 bp. Reads with Q-scores < 10 were excluded from further analysis. The EPI2ME pipeline includes Fastcat for quality assessment, Kraken2 for taxonomic classification, and TaxonKit for annotation refinement. Alpha diversity metrics (Shannon, Simpson, and Chao1 indices) were calculated using EPI2ME v5.2.5. Taxonomic profiles were analyzed at the phylum, class, order, family, genus, and species levels. Predicted functional metagenomic profiling was conducted using Phylogenetic Investigation of Communities by Reconstruction of Unobserved States (PICRUSt2) (Douglas et al. 2020). Functional pathways were annotated against the Kyoto Encyclopedia of Genes and Genomes (KEGG) and MetaCyc databases (Kanehisa 2002; Karp et al. 2002). Dietary intake data obtained from the SQ-FFQ were analyzed using GraphPad Prism software (version 10.6, GraphPad Software, USA). Depending on data distribution, appropriate parametric or nonparametric tests were applied. A p-value < 0.05 was considered statistically significant.

## Results And Discussion

### Participant Characteristics and Anthropometric Profiles

A total of 36 children aged 2–5 years were enrolled, consisting of 18 stunted and 18 normal participants. The mean age and sex distribution were comparable between groups, ensuring a balanced cohort for microbial comparison. Anthropometric measurements demonstrated significant differences in all nutritional indices. The mean height-for-age Z-score (HAZ) was markedly lower in the stunted group (−2.6 ± 0.4) than in the normal-weight group (0.5 ± 0.7; P < 0.0001). Similarly, weight-for-age (WAZ) and weight-for-height (WHZ) were both significantly reduced in stunted children (P < 0.0001), confirming the phenotypic classification and nutritional deficit associated with growth retardation. These anthropometric contrasts established a clear clinical basis for assessing microbial and metabolic disparities between the two groups.

### Gut Microbial Richness and Alpha Diversity

Alpha diversity indices revealed a consistent reduction in gut microbial richness and evenness among stunted children. The Chao1 richness index (Figure 1A) appeared lower in the stunted pooled sample compared with the normal pooled sample, indicating reduced estimated microbial richness. Correspondingly, Shannon and Simpson diversity indices (Figure 1B) showed a trend toward reduced diversity, suggesting not only fewer species but also an uneven microbial distribution within the community. The evenness index further confirmed dominance by a limited number of bacterial taxa in stunted children, consistent with an unstable and dysbiotic gut environment. This reduction in microbial diversity implies a weakened ecological resilience of the intestinal microbiota, which could compromise the community’s ability to perform critical metabolic and immunomodulatory functions.

**Figure 1.**
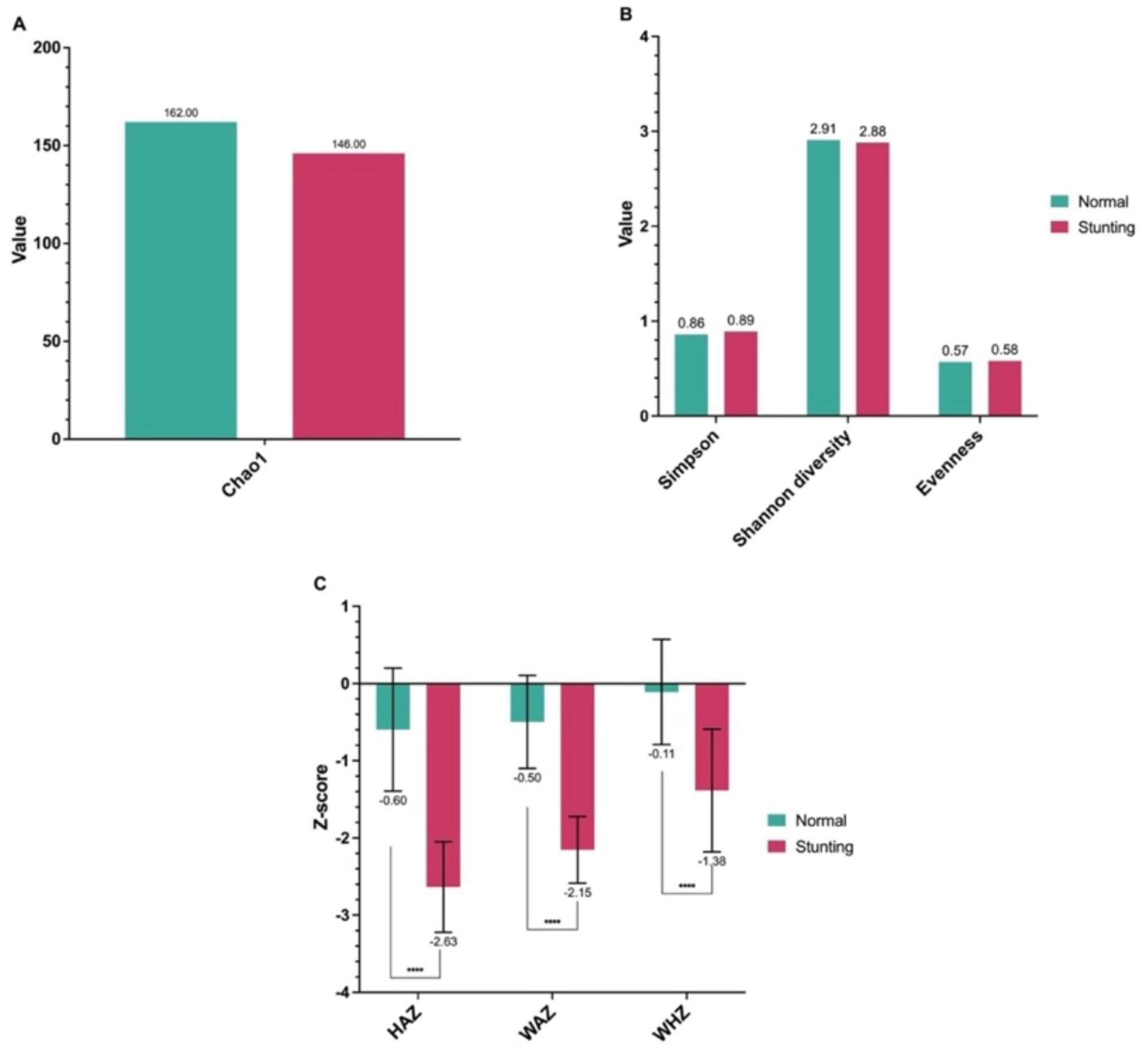
Comparison of gut microbiota diversity indices and anthropometric Z-scores between normal and stunting children. (A) Chao1 richness index, (B) alpha diversity indices including Simpson, Shannon diversity, and Evenness, and (C) anthropometric Z-scores for height-for-age (HAZ), weight-for-age (WAZ), and weight-for-height (WHZ). Data are presented as mean values ± standard deviation. Significant differences between groups are indicated (**** p < 0.0001).

### Taxonomic Composition of Gut Microbiota

The taxonomic profiling of the gut microbiota revealed clear compositional distinctions between stunted and normal children across multiple phylogenetic levels (Figure 2). At the phylum level, both groups were overwhelmingly dominated by Bacillota which accounted for more than 96% of total reads in each group indicating a Bacillota rich microbial ecosystem in this population. Although Bacillota remained the dominant phylum in both groups, its proportion was slightly higher in stunted children (96.54%) than in normal children (96.04%). In contrast, Pseudomonadota exhibited a higher relative abundance in normal children (3.22%) compared with stunted children (2.40%). Bacteroidota was also more abundant in stunted children (0.60%) relative to normal children (0.30%) suggesting limited expansion of this fiber degrading phylum. Actinomycetota appeared in low proportions in both groups and showed a minor decrease in stunted children. Synergistota was detected only in the normal group, whereas Verrucomicrobiota appeared exclusively in the stunted group although at extremely low abundances.

**Figure 2.**
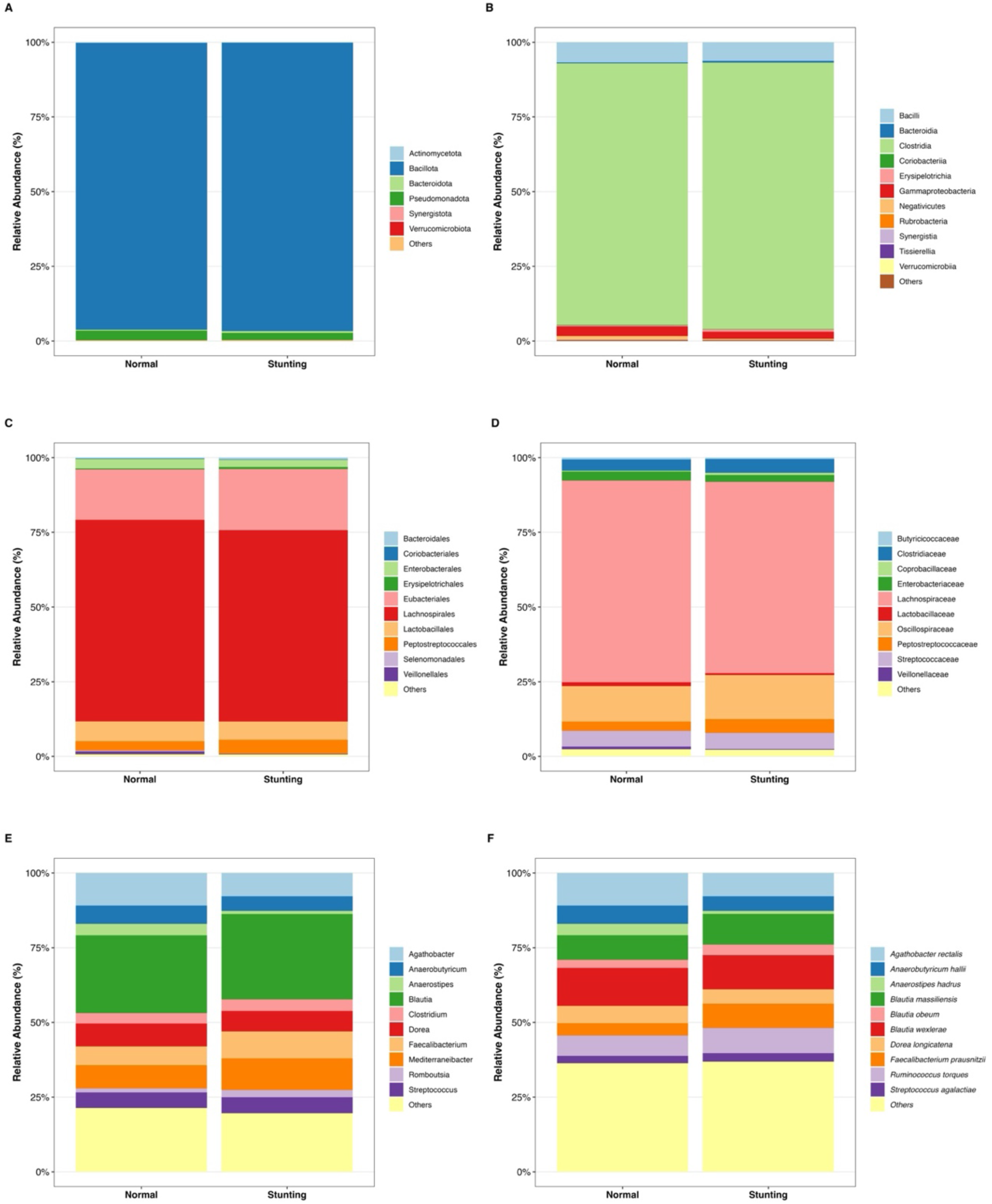
Relative abundance of gut microbiota composition between normal and stunting children at different taxonomic levels. (A) Phylum level, (B) Class level, (C) Order level, (D) Family level, (E) Genus level, and (F) Species level. Each stacked bar represents the mean relative abundance (%) of dominant taxa in each group. Differences in microbial community structure between normal and stunting groups are visualized by variations in color-coded taxa proportions.

At the class level (Figure 2B), Clostridia overwhelmingly dominated the gut microbiota in both groups accounting for 87.52% of sequences in normal children and increasing to 89.17% in stunted children. This slight elevation in stunted children reflects a further shift toward Bacillota associated anaerobic fermenters. Bacilli represented the second highest class with 6.76% in normal children and 6.22% in stunted children. Although the difference was small, the reduction in stunted children may indicate altered carbohydrate fermenting activity. Among minority classes, Bacteroidia increased from 0.30% in normal children to 0.60% in stunted children, while Gammaproteobacteria decreased from 3.22% to 2.40% mirroring patterns observed at the phylum level. Notably, Erysipelotrichia nearly doubled in stunted children (0.76% vs 0.35%) consistent with associations between this class and inflammation or impaired metabolic function. Other low abundance classes showed minimal but distinct patterns such as reduced Negativicutes in stunted children (0.38% vs 1.30%) and the exclusive presence of Verrucomicrobiae in stunted children at very low levels (0.01%).

At the order level (Figure 2C), the microbiota of both groups was dominated by Lachnospirales comprising 67.44% of reads in normal children and 64.02% in stunted children. This slight reduction in stunted children indicates a modest loss of fiber fermenting taxa. Eubacteriales another major Bacillota order increased notably in stunted children (20.42% vs 16.88%) suggesting a shift toward Clostridia associated fermentative and potentially pro inflammatory taxa. In contrast, Enterobacterales the main order within Gammaproteobacteria decreased from 3.15% in normal children to 2.38% in stunted children. Meanwhile, Peptostreptococcales increased from 3.11% to 4.66% reflecting enrichment of taxa frequently associated with dysbiosis. Less abundant orders including Erysipelotrichales and Acidaminococcales also showed higher proportions in stunted children indicating perturbations in fermentation and amino acid metabolism.

At the family level (Figure 2D), the most substantial shift occurred in Oscillospiraceae rising from 11.87% in normal children to 14.72% in stunted children suggesting altered butyrate production dynamics. Meanwhile, Lachnospiraceae the dominant family across both groups decreased slightly in stunted children (64.02% vs 67.44%). In the Bacteroidota phylum, Prevotellaceae showed a marked increase in stunted children (0.60% vs 0.13%) while Bacteroidaceae were present only in normal children (0.17%). Conversely, Enterobacteriaceae a key inflammation associated family decreased from 3.06% to 2.34%. Families connected to inflammatory or dysbiotic states such as Clostridiaceae Peptostreptococcaceae and Erysipelotrichaceae exhibited higher relative abundance in stunted children.

At the genus and species level, the gut microbiota of both groups was dominated by members of the Lachnospiraceae and Oscillospiraceae families yet several clear divergences appeared between normal and stunted children. Normal children showed higher relative abundance of beneficial butyrate producers such as *Agathobacter rectalis* (10.86% vs 7.79%), *Anaerobutyricum hallii* (6.18% vs 4.92%), *Dorea longicatena* (5.75% vs 4.79%), and *Coprococcus eutactus* (0.82% vs 0.81%). In contrast, stunted children exhibited enrichment of several fermentative or dysbiosis associated taxa particularly Blautia species including *Blautia massiliensis* (10.32% vs 8.31%), *Blautia obeum* (3.53 % vs 2.70 %), and *Blautia wexlerae* (11.42 % vs 12.70 %) along with higher levels of Romboutsia species and *Terrisporobacter petrolearius* (0.23 % vs 0.06 %). Several opportunistic species appeared more frequently in stunted children including *Streptococcus agalactiae*, *Streptococcus pneumoniae*, and the mucin degrader *Akkermansia muciniphila* which was detected only in the stunted group. Conversely, *Escherichia coli* and other Enterobacteriaceae taxa such as Citrobacter and Klebsiella were slightly higher in normal children reflecting the overall reduction in Gammaproteobacteria in stunting. Short chain fatty acid producing *Faecalibacterium prausnitzii* increased substantially in stunted children (8.17 % vs 4.20 %) suggesting compensatory shifts within Oscillospiraceae while species indicative of disturbed protein fermentation such as *Clostridium perfringens*, *Clostridium septicum*, and *Clostridioides difficile* were also more abundant in stunted children. Collectively, stunted children displayed a gut microbial profile characterized by reduced diversity of health associated butyrate producers alongside enrichment of fermentative mucin degrading and dysbiosis linked species indicating a shift toward a metabolically imbalanced gut ecosystem.

### Predicted Functional Metabolic Pathways

Predicted functional profiling using PICRUSt2 revealed marked differences in microbial metabolic potential between normal and stunted children. KEGG pathway analysis (Figure 3A) showed that stunted children exhibited higher predicted abundances across multiple metabolic categories, including carbohydrate, amino acid, nucleotide and energy metabolism. Pathways such as starch and sucrose metabolism, porphyrin metabolism, pyrimidine metabolism and peptidoglycan biosynthesis appeared more abundant in stunted children, suggesting increased microbial metabolic turnover. Similarly, MetaCyc functional predictions (Figure 3B) indicated higher representation of carbohydrate-processing pathways in the stunted group, including sucrose biosynthesis II, starch degradation III, glycogen biosynthesis I and glycolysis-related pathways.

**Figure 3.**
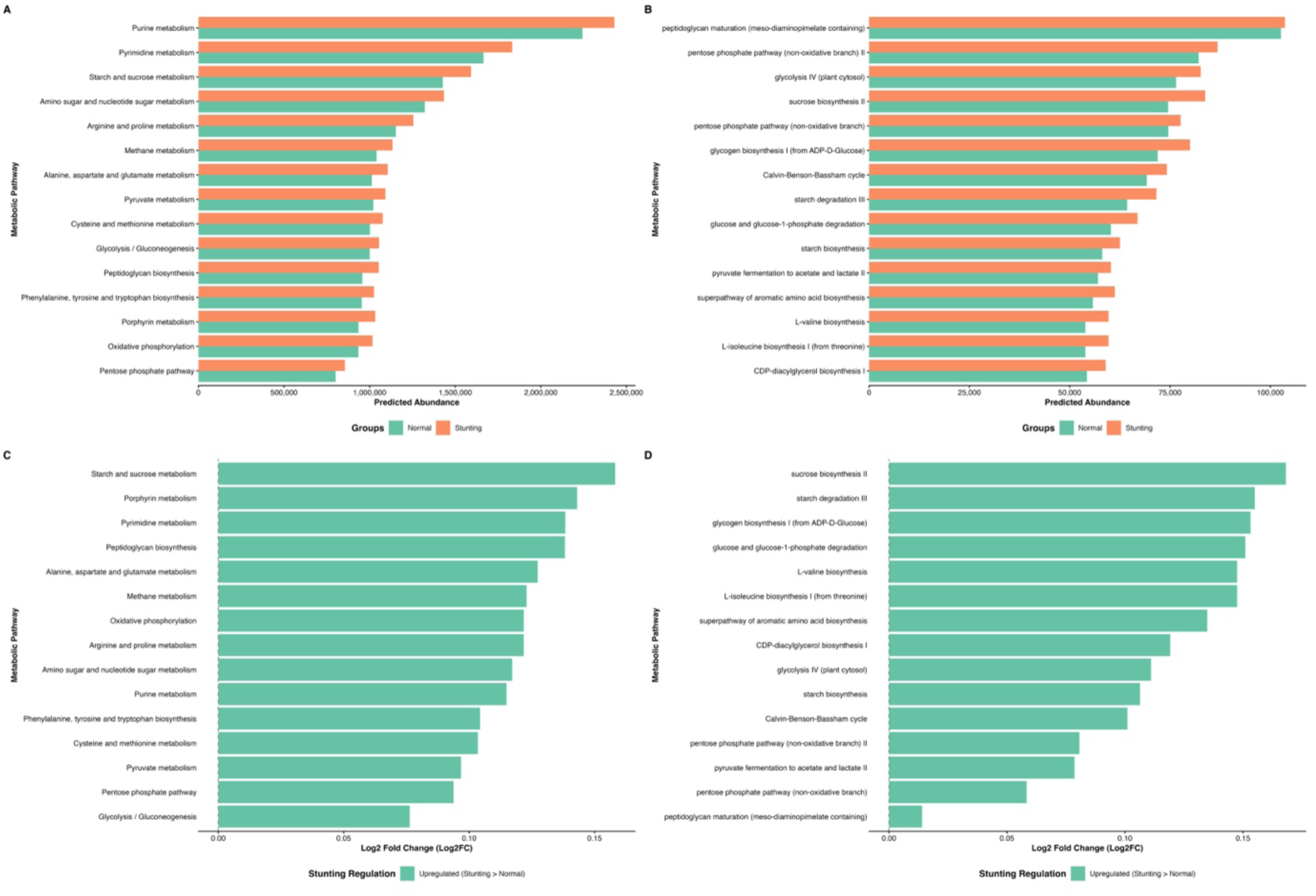
Predicted functional profiling of gut microbiota between normal and stunting children based on KEGG metabolism pathway analysis and METACYC. (A) Comparison of top predicted metabolic pathways at KEGG, (B) top predicted pathways at METACYC, (C) upregulated KEGG metabolic pathways in the stunting group (Log₂ fold change), and (D) upregulated METACYC pathways in the stunting group (Log₂ fold change). The predicted abundances were inferred using PICRUSt2 analysis. Green bars represent the normal group, while orange bars represent the stunting group.

**Figure 4.**
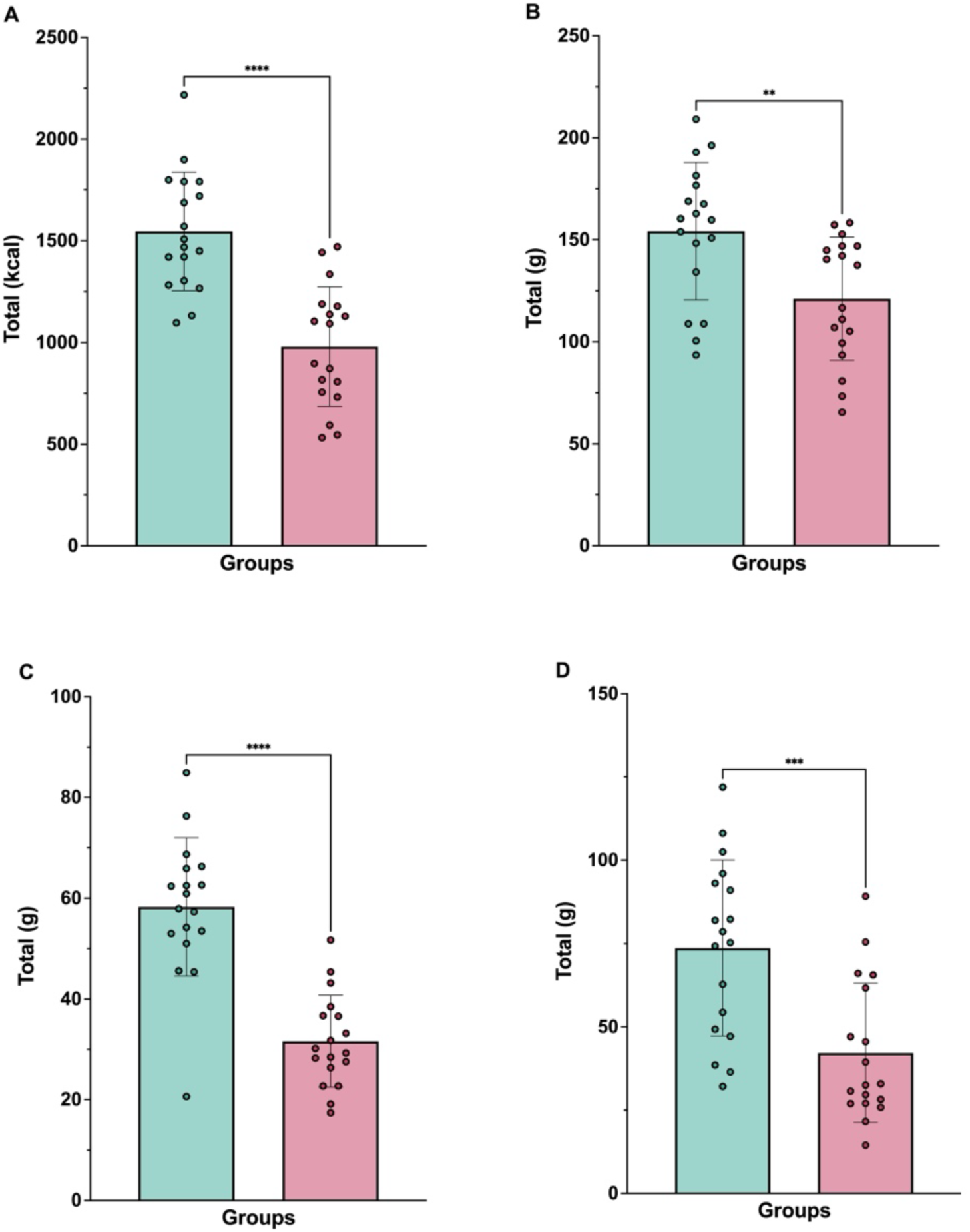
Comparison of macronutrient intake between normal and stunting children based on the Semi-Quantitative Food Frequency Questionnaire (SQ-FFQ). (A) Total energy intake (kcal), (B) carbohydrate intake (g), (C) protein intake (g), and (D) lipid intake (g). Data are presented as mean ± standard deviation. Statistical differences between groups were determined using appropriate tests, with significance indicated as p** < 0.01**, \*\*\***p < 0.001, and **\*\*\*\***p < 0.0001.

The magnitude of these functional differences is illustrated in the Log₂ Fold Change plots (Figure 3C–D). KEGG functions with the largest positive Log₂FC values in the stunted group included starch and sucrose metabolism (Log₂FC = 0.158), porphyrin metabolism (0.143), pyrimidine metabolism (0.138), and alanine, aspartate and glutamate metabolism (0.127). MetaCyc pathways showed similar trends, with the highest Log₂FC values observed for sucrose biosynthesis II (0.168), starch degradation III (0.155), and glycogen biosynthesis I (0.153). These findings indicate broad enhancement of predicted carbohydrate breakdown, amino acid biosynthesis and energy production pathways in stunted children.

Collectively, the data presented in Figure 3A–D point to a functional shift in the gut microbiota of stunted children toward greater fermentative capacity, increased biosynthetic activity and elevated metabolic flux, patterns that may contribute to intestinal stress, increased nutrient demand and impaired gut barrier function. These functional features are consistent with a dysbiotic microbial configuration and may underlie the physiological disturbances associated with stunting.

### Macronutrient Intake and Its Association with Microbiota

Analysis of dietary intake revealed a clear nutritional disparity between stunted and normal-weight children. Total energy intake was lower in the stunted group, accompanied by significant reductions in macronutrient consumption. Protein intake was markedly lower (***P < 0.001), while lipid intake showed an even greater decline (****P < 0.0001). Carbohydrate intake, though reduced, did not reach the same level of statistical difference. These macronutrient deficiencies may contribute to impaired microbial substrate availability, leading to a decline in beneficial fermentative bacteria. The insufficient intake of proteins and fats likely exacerbates the functional limitations of the stunted microbiota, reducing its capacity for SCFA production and metabolic balance.

### Micronutrient Intake Profiles and Correlation with Gut Microbiota

Micronutrient assessment indicated consistently lower intakes among stunted children (Figure 5A–L). Vitamins B1, B2, B6, and B9 (folate) were significantly lower (P < 0.05–P < 0.001), reflecting inadequate dietary intake of key cofactors essential for both host and microbial metabolism. Reduced intake of vitamin B6 and folate aligns with the predicted depletion of microbial pathways responsible for their biosynthesis (Figure 3A–B), suggesting a synergistic deficiency at both the host and microbial levels. Additionally, essential minerals such as calcium, magnesium, iron, and zinc were significantly reduced (P < 0.01–P < 0.001). These micronutrients play critical roles in immune function, energy metabolism, and microbial enzymatic activity. Their deficiency may not only impair host physiological processes but also further destabilize microbial community composition, perpetuating the cycle of malnutrition and dysbiosis.

**Figure 5.**
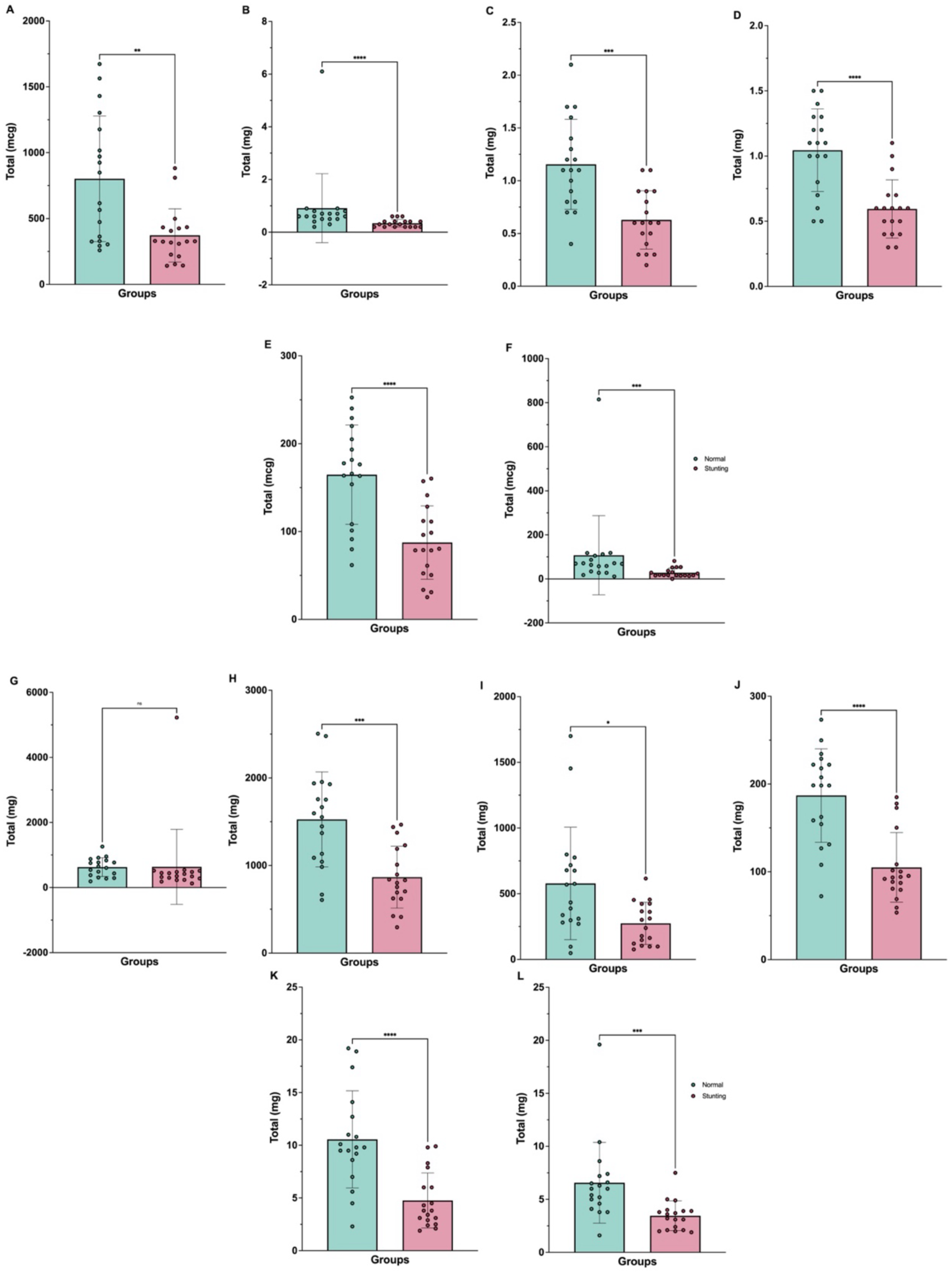
Comparison of micronutrient intake between normal and stunting children based on the Semi-Quantitative Food Frequency Questionnaire (SQ-FFQ). (A) Vitamin A, (B) Vitamin B₁, (C) Vitamin B₂, (D) Vitamin B₆, (E) Vitamin B₉, (F) Vitamin C, (G) Sodium, (H) Potassium, (I) Calcium, (J) Magnesium, (K) Iron, and (L) Zinc. Data are presented as mean ± standard deviation. Statistical differences between groups were determined using appropriate tests, with significance indicated as p^ns^ **≥** 0.05, p* < 0.05, p** < 0.01**, \*\*\***p < 0.001, and **\*\*\*\***p < 0.0001.

## Discussion

This study provides the first characterization of the gut microbiota of stunted children in Banyumas Regency, a region with persistent nutritional and environmental challenges. Using full-length 16S rRNA sequencing on pooled fecal samples, we identified distinct microbial and functional patterns that differentiate stunted children from their normal-weight peers. These findings illustrate a dysbiotic configuration that, although sharing some features with previously reported stunting-associated microbiomes, also displays several unique signatures specific to this population. Consistent with earlier literature, stunted children exhibited lower alpha diversity, reflected by reduced richness and evenness. Reduced microbial diversity has been linked to ecological instability, diminished metabolic capacity, and impaired resilience to environmental perturbation (Chibuye et al. 2024; Khan Mirzaei et al. 2020) Such simplified microbiome structures are frequently observed in malnourished children and are considered markers of an “immature” microbiome state that fails to support optimal nutrient metabolism and immune maturation (Subramanian et al. 2014)

Taxonomic profiling revealed patterns that differ in notable ways from previously published work. Both groups were overwhelmingly dominated by Bacillota (>96%), a finding consistent with populations consuming high-carbohydrate diets and reflecting a strongly fermentative gut environment (De Filippo et al. 2010). However, subtle but meaningful differences emerged between groups. Stunted children exhibited a modest increase in Bacillota and a reduction in Pseudomonadota compared with normal-weight children. This pattern contrasts with many studies in Bangladesh, Malawi, and Indonesia reporting elevated Proteobacteria, including Enterobacteriaceae as a hallmark of stunting-associated dysbiosis (Masrul et al. 2020; Smith et al. 2013; Surono et al. 2021). These findings suggest that dysbiosis in Banyumas may follow a different ecological trajectory. At finer taxonomic resolution, stunted children showed increased abundance of several Clostridia-affiliated orders and families, including Eubacteriales, Peptostreptococcales, Clostridiaceae, and Erysipelotrichaceae groups frequently associated with mucosal inflammation, amino acid fermentation, and metabolic stress (Kaakoush 2015; Kaakoush et al. 2014; Lopetuso et al. 2023). In contrast, taxa linked to fiber degradation and mucosal homeostasis, such as Lachnospiraceae, were proportionally higher in normal children, consistent with their roles as key producers of short-chain fatty acids (SCFAs) essential for epithelial integrity and anti-inflammatory signaling (Louis and Flint 2017).

Interestingly, several SCFA-producing taxa, including *Faecalibacterium prausnitzii* and multiple *Blautia* species, were more abundant in stunted children. Although counterintuitive, similar compensatory increases in SCFA producers have been reported in other undernourished cohorts and may reflect altered energy extraction demands or increased turnover of luminal substrates (Blanton et al. 2016). Importantly, greater microbial abundance does not guarantee functional adequacy; impaired epithelial uptake of SCFAs has been repeatedly documented in stunted children with environmental enteric dysfunction (EED) (Harper et al. 2018). Genus and species-level transitions further support a shift toward fermentative and dysbiosis-associated taxa. Enrichment of *Blautia massiliensis*, *Blautia wexlerae*, *Romboutsia* spp., and *Terrisporobacter petrolearius* points to increased protein fermentation and redox imbalance in the gut features linked to impaired growth and intestinal inflammation (Chen et al. 2020). The presence of opportunistic taxa such as *Streptococcus agalactiae*, *Streptococcus pneumoniae*, and *Akkermansia muciniphila* in the stunted group suggests mucosal barrier disruption and enhanced mucin turnover, common features of EED (Derrien et al. 2017; Vonaesch et al. 2018). In contrast to many global reports, Enterobacterales were slightly higher in normal children in this cohort, reinforcing the regional specificity of microbiome profiles.

Functional predictions provided additional insight into microbial metabolic behavior. KEGG and MetaCyc analyses revealed that stunted children exhibited higher predicted abundances across carbohydrate, amino acid, nucleotide, and energy metabolism pathways. Enhanced representation of starch and sucrose metabolism, glycolytic pathways, alanine/glutamate metabolism, porphyrin metabolism, and nucleotide biosynthesis may reflect increased microbial metabolic turnover and heightened competition with the host for limited substrates (Rooks and Garrett 2016). This hyperactive yet potentially inefficient metabolic profile contrasts with some studies reporting functional depletion in stunted microbiomes, highlighting population-specific variations in metabolic signatures.

Dietary intake patterns offer important context for these findings. Stunted children consumed significantly less energy, protein, fat, and multiple micronutrients especially B vitamins and essential minerals. Dietary deficiencies are known to alter microbial substrate availability, suppress saccharolytic taxa, and promote enrichment of amino acid–fermenting or mucin-degrading bacteria (Muramatsu and Winter 2024). Reduced intake of B vitamins aligns with the increased microbial biosynthetic pathways predicted in the stunted group, suggesting potential microbial compensation for host nutrient deficiency. Meanwhile, low fiber intake may partially explain reduced proportions of Lachnospiraceae and other beneficial fiber-degrading taxa. Collectively, the taxonomic and functional signatures observed here support a model in which stunted children in Banyumas harbor a dysbiotic microbiota characterized not by the canonical Proteobacteria expansion described in many regions, but by an altered fermentative ecosystem driven by Clostridia-dominated community shifts, increased metabolic flux, and enrichment of taxa associated with mucosal stress. These microbial features, combined with inadequate nutrient intake, may contribute to impaired barrier function, inefficient nutrient assimilation, and chronic low-grade inflammation, which are key drivers of growth faltering and EED pathophysiology (Amimo et al. 2025).

This study has limitations. Pooled samples restrict assessment of inter-individual variability and limit statistical comparisons. Functional predictions are based on 16S rRNA inference and should be validated with shotgun metagenomics or metabolomics. Nonetheless, the data establish an important microbial baseline for children in Banyumas and reveal unique ecological signatures that merit further investigation. Future research incorporating longitudinal sampling, metagenomic functional profiling, and dietary interventions is needed to elucidate causal mechanisms linking dysbiosis, nutrition, and growth. The present findings underscore the importance of developing microbiota-informed public health strategies tailored to the ecological context of Indonesian children.

## Data Availability

All data produced in the present work are contained in the manuscript

## Acknowledgements

The authors thank the Institute for Research and Community Service (LPPM), Universitas Jenderal Soedirman, for funding this study through research grant scheme in 2025 with decree No. 14.539/UN23.34/PT.01/V/2025. The authors gratefully acknowledge this support, which made the implementation of this research possible.

